# Multidimensional motivation in aging: a validated framework spanning goal-directed behaviour, social reward and pleasure

**DOI:** 10.64898/2026.06.11.26355497

**Authors:** Samuel L. Warren, Haridra Somasundaram, Kristina Horne, Gail A. Robinson, Julie Behenská, Kathy Nguyen, Pranav Ajay, Anita M.Y. Goh, Yun-Hee Jeon, Lee-Fay Low, Chang Xu, Muireann Irish, the MotDem Consortium

**Author notes:** **MotDem Consortium:** Rebekah M. Ahmed, Isabelle Burke, Jade Cartwright, Tao Chen, Trevor Chong, Sally Day, David Foxe, Fiona Kumfor, Samantha Loi, Olivier Piguet, Siobhán Shaw, Dennis Velakoulis, Alexis E. Whitton, Stephanie Wong (see Appendix for full details). Corresponding author: Professor Muireann Irish, Brain and Mind Centre, The University of Sydney, NSW 2050, Australia.

## Abstract

Motivational changes are determinants of healthy aging, social engagement, and functional independence, and may signal early neurodegenerative risk. Existing assessment approaches in aging typically treat motivation as a unitary construct. Here, we introduce MotDem, an age-appropriate measure of motivation co-designed with people living with dementia, carers, and clinicians. Across a broad adult lifespan sample (18-80 years), MotDem revealed a robust three-domain motivational architecture encompassing goal-directed behaviour, social reward, and pleasure, with a fourth satiety factor retained as exploratory. This structure was replicated in an independent older cohort (45-80 years) from a different national context. MotDem showed strong convergence with established measures of apathy and anhedonia, alongside more modest associations with depressive symptomatology. Together, these findings show that motivational aging is multifaceted and poorly captured by traditional unitary assessment. MotDem provides a multidimensional framework for measuring distinct motivational drivers of heterogeneous aging trajectories, with implications for resilience, wellbeing, and neurodegenerative risk.

## Introduction

Motivation is a fundamental driver of behaviour across the lifespan, influencing how individuals initiate, sustain and derive meaning from everyday activities. In later life, motivational processes are increasingly recognised as key determinants of healthy aging, influencing social participation, functional independence and overall quality of life. Age-related changes in motivation and reward-processing can influence engagement in physical, social, and cognitive activities, with downstream implications for brain health trajectories and wellbeing.^1,2^ Importantly, accumulating evidence suggests that alterations in motivation may precede overt cognitive or functional decline, suggesting motivation may be a potentially sensitive indicator of neurodegenerative risk.^3–5^

Motivational shifts in later life are not inherently pathological and may reflect adaptive responses to changing life contexts, resources, and priorities. Motivation is a complex and multidimensional construct, broadly encompassing the capacity to initiate and sustain goal-directed behaviour, sensitivity to social rewards, and the experience of pleasure. These dimensions are unlikely to be homogenous or to follow a single trajectory across ages, showing distinct patterns of change in later life. Importantly, these dimensions span discrete motivational processes rather than representing subdomains within a single behavioural construct. Alterations in these dimensions may reflect age-related changes in cognitive function alongside a recalibration of goals and values.^6–8^ Crucially, these dimensions may not carry equal weighting in terms of later-life outcomes.^1^ For example, alterations in goal-initiation or effort mobilisation may disproportionately affect health-related behaviours such as physical activity and functional independence,^8^ whereas changes in reward sensitivity or social motivation may have more pronounced consequences for emotional wellbeing, social engagement, and longevity.^9,10^ Disruption of specific motivational components may therefore contribute to divergent aging trajectories and risk profiles in later life.

Characterising motivational changes in aging requires a multidimensional approach that can distinguish core motivational components, track their trajectories across adulthood, and clarify their differential relevance for functioning and wellbeing in later life. Current research approaches to assessing motivational changes in later life remain limited, with many studies using subscales or item combinations extracted from depression screening tools.^1,11^ These measures are valuable in that they capture changes in mood that are concurrently associated with loss of motivation, cognitive decline, and increased dementia risk. For example, accumulating evidence suggests that age-related conditions including neuropsychiatric syndromes (e.g., late-life depression) and dementia are often associated with reduced motivation to engage in effortful activities.^12^ However, existing depression-based measures tend to prioritise the hedonic elements of anhedonia while neglecting motivational components of this symptom. As such, measures of motivational impairment derived from depression screening scales will provide at best an indirect or unidimensional measure of such constructs.

In the same vein, measures of apathy have proven highly valuable for screening for motivational changes later in life. Many widely used apathy instruments, however, do not capture several motivational dimensions that are relevant for older people,^1,13^ such as social reward processing and the hedonic experience of pleasure and enjoyment.^14^ Changes in these dimensions may occur independently of low mood and can follow trajectories that are distinct from affective symptomatology.^15^ As such, age-related reductions in reward sensitivity or a narrowing of social motivation may meaningfully constrain participation in physical, cognitive, and social activities even when depressive symptoms are minimal.^16^

Converging evidence from large-scale population studies indicates that both social engagement and hedonic tone show strong predictive utility for long-term health outcomes including longevity.^9^ Positive enjoyment of life, in particular, has been shown to predict survival even when accounting for pre-existing illness, health behaviours such as smoking and physical activity, and depressive symptoms.^10^ In this context, pleasure (i.e., anticipatory or consummatory reward) emerges as a distinct motivational construct with substantial protective effects and independent associations with health outcomes in later life.^10^ Collectively, these findings indicate that assessment of motivational changes in aging requires explicit consideration of pleasure and social reward alongside more traditional indices of goal-directed behaviour. Failure to index these discrete motivational dimensions limits our ability to model how motivation changes across the lifespan and to characterise key pathways through which motivation influences engagement, wellbeing, and health trajectories in older age.

To address these limitations, we propose a framework that conceptualises motivation in aging as a multidimensional construct with distinct components that may change across the lifespan. This framework differentiates core dimensions of motivation that hold particular relevance in later life, namely the capacity to initiate and sustain goal-directed behaviour, sensitivity to social reward, and the consummatory experience of pleasure and enjoyment. Together these dimensions align with the positive valence domain of the Research Domain Criteria (RDoC) framework^17^ and capture complementary motivational processes that influence engagement, wellbeing, and aging trajectories.

The present study aimed to operationalise a multidimensional framework of motivation in aging through the development of MotDem (i.e., Motivation in Aging and Dementia), a brief, age-appropriate measure suitable for use across the adult lifespan and in neurodegenerative populations. Specifically, we sought to (i) identify core domains of motivational functioning in aging guided by lived experience and expert input, (ii) develop and refine a measure of these domains through an embedded co-design process involving older adults, people living with dementia, and current and former carers, (iii) evaluate the psychometric properties and construct validity of this framework in a large adult lifespan sample, and (iv) examine its generalisability in an independent cohort from a different national context. In addition, we sought to provide age-adjusted normative data to support interpretation across the adult lifespan.

In doing so, we aim to move beyond unitary and mood-centred models of motivation and to establish a framework capable of capturing distinct motivational processes that shape heterogeneous aging trajectories.

## Methods

The following sections describe three methodological stages that map directly onto the aims outlined above. Stage 1 focuses on questionnaire development, including systematic review of existing measures, iterative item refinement, and co-design with people with lived experience and relevant professionals. Stage 2 comprises refinement and validation of the instrument, including recruitment of two independent cohorts (discovery and external validation samples) and associated statistical analyses. Stage 3 details the development of the normative MotDem model and derivation of age-adjusted reference values.

## Participants

### Discovery cohort

An initial discovery cohort of 421 participants was recruited through Amazon Mechanical Turk (MTurk) using CloudResearch^18^ for the purpose of questionnaire validation (Stage 2). Recruitment was stratified across three age groups (18–40, 41–59, and >60 years) with approximately 140 participants sampled per group. This approach ensured balanced age representation during recruitment and was optimised using CloudResearch’s Approved Participants feature.^18^ All participants were residents of the United States of America and were monetarily reimbursed for their time ($6.00 USD). Participant responses were screened by the research team, with any indication of automated completion (i.e., a script or bot) removed (e.g., electronic signatures inserted into clock drawing space for ACE-III; n = 27). An additional 40 participants were excluded due to self-reported history of major psychiatric or neurological condition, significant acquired brain injury, or current use of medications that can significantly affect cognition (e.g., antipsychotics, benzodiazepines, anti-convulsants). The final sample included 354 participants (see Table 1 for descriptive statistics by age group). The study was approved by the University of Sydney local Ethics Committee (HREC: 2023/655). All participants provided informed consent in accordance with the Declaration of Helsinki.

**Table 1.**
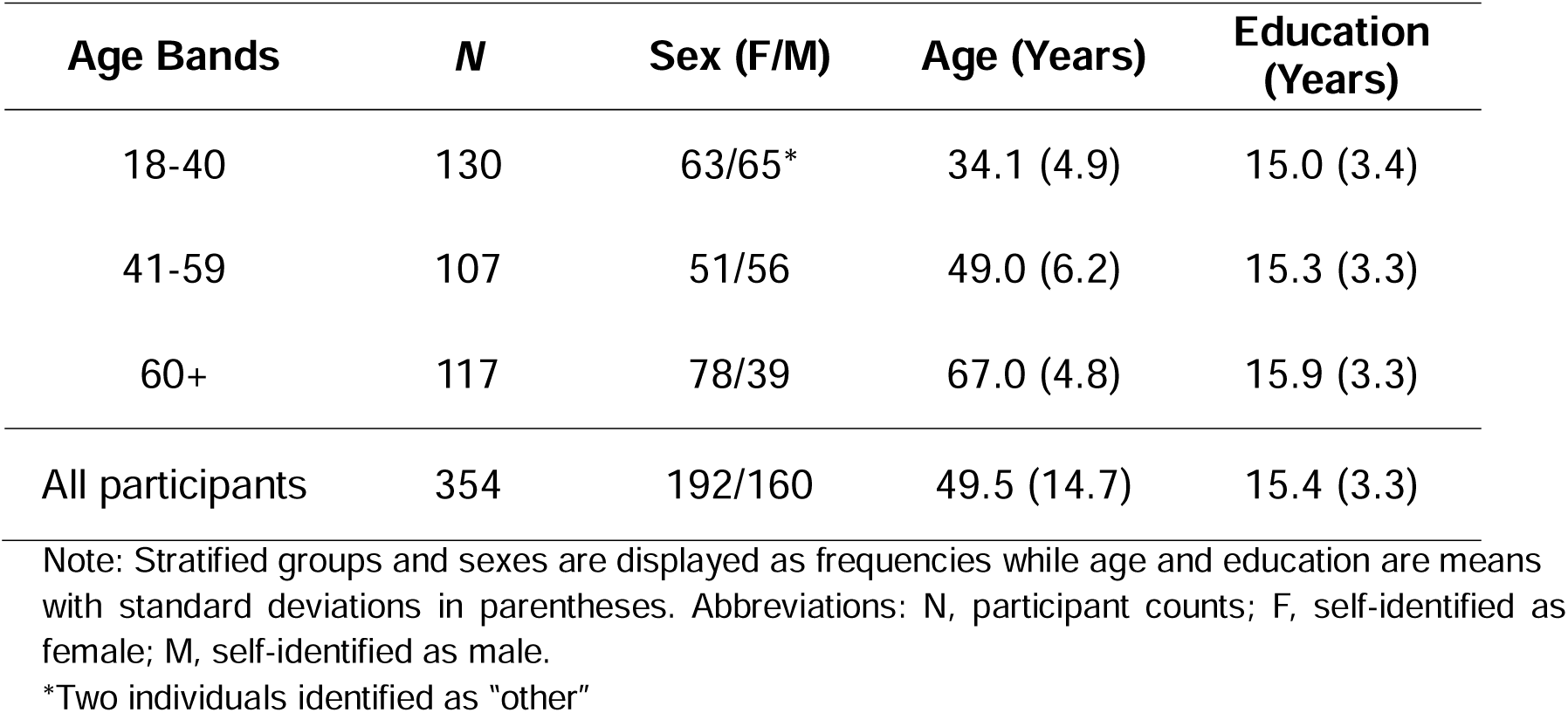
Descriptive statistics for each age-stratified recruitment wave of the discovery cohort.

## External validation cohort

To evaluate the robustness and cross-national generalisability of MotDem (Stage 2), we recruited an independent Australian sample of healthy middle-to-older adults aged 45-80 years (*N*=241; *M* = 64.4, *SD* = 7.6) through the StepUp for Dementia research participation and engagement platform.^19^ This cohort provided a targeted validation sample distinct from the discovery sample in terms of national setting, age distribution, and recruitment pathway, enabling us to assess the stability of MotDem motivational dimensions in a mid-to-later life population for whom motivational screening is most relevant. MotDem Total and subscale scores were examined for profile stability and consistency with the discovery sample.

## Procedures

### Systematic review of existing tests

In Stage 1, a systematic review was undertaken in PsycTests (Ovid) and PubMed to identify relevant psychometric scales. Our search criterion was limited to articles from January 1990 to the search date (August 2023), studies conducted in humans, and articles published in English. When translations or adaptations of a scale existed, the most appropriate version was selected. Four separate searches were performed to ensure all possible scales measuring motivation/apathy and pleasure/anhedonia were captured. The first three searches were performed in PsycTests for each specific construct separately, i.e., anhedonia.mp, apathy.mp, motivation.mp (where mp=test name, test description, heading word, key concepts). The fourth search was performed in PubMed using the following keywords; “motivation”, “apathy”, “anhedonia”, “effort”, “pleasure”, AND “assessment”, “test”, “questionnaire”, “measure, “scale”.

The systematic review search was conducted by two raters (MI, AS), who independently reviewed all search results and identified 50 unique scales assessing dimensions of motivation and pleasure. Questionnaires were included if the overall scale, or one or more subscales, specifically targeted at least one of the core constructs of motivation/apathy, pleasure/anhedonia and used a Likert scale/rating approach for responses. See Supplementary Table 1 for the full list of extracted questionnaires measuring apathy and anhedonia that were reviewed during MotDem item development.

### Item extraction and prioritisation

Items were extracted from each questionnaire for review. Where instruments captured multiple constructs, only items explicitly assessing motivation/apathy and pleasure/anhedonia or related processes (e.g., reward sensitivity, effort expenditure) were retained. This process yielded an initial pool of 1325 items. Next, items were independently screened by two authors (MI and AS) to remove duplicates and exclude items that fell outside the intended scope of MotDem, including those assessing personality traits, open-ended or interview-based responses, ecological momentary assessment formats, or extended temporal periods (>1 month). Through iterative screening, the item pool was reduced to 334 unique items for further evaluation.

Remaining items were standardised through rewording into first-person format, deidentified from their source measures, and provisionally organised into candidate domains reflecting key aspects of motivation and reward processing (e.g., anticipatory and consummatory pleasure, effort/persistence, reward sensitivity, satisfaction, savouring) and reward type (e.g., food/drink, hobbies/interests, intimacy, prosocial, social, sensory).

### Panel ratings

A panel of five raters from the broader research team comprising clinical neuropsychologists, a clinical psychologist, a neuroscientist, and a neurologist independently evaluated each item for inclusion. Items were assessed based on how well they captured core aspects of motivation, reward processing, and pleasure, as well as their clarity and relevance for real-world manifestations of motivation change.

To ensure applicability across normative and clinical contexts, raters were instructed to consider the extent to which items reflected motivational changes observed in healthy aging and neurodegenerative conditions such as dementia. This approach was intended to maximise generalisability while maintaining sensitivity to clinically meaningful variation in motivation.

Raters evaluated items within each thematic category and indicated whether each item should be retained at this stage. Items were endorsed for inclusion if they were judged to capture relevant constructs and/or could be refined or combined with related items. Items were excluded if they were considered ambiguous, difficult to interpret, not directly related to the constructs of interest, or better captured by existing measures.

Items assigned a “Yes” response were given a value of 1, while items with a “No” response were given a value of 0. This enabled us to calculate the variance across raters as a natural metric to measure item divergence. Of the 334 items, 53 showed complete concordance (i.e., agreement by all 5 raters, 0 variance). From these, 3 items were removed as raters had unanimously endorsed a “No” response. Next, we reviewed “Yes” items where 4 of the 5 raters showed agreement (i.e., 0.2 variance) and retained 26 items that indexed the domains of effort and initiation, as these were deemed important to include in the context of early diagnosis of dementia. As a final robustness check, we ran a matrix factorisation-based approach (non-negative matrix factorisation implemented in scikit-learn) to the annotation data to obtain a low-rank approximation and minimise the influence of individual rater variability. Following this procedure, we re-sorted the variance of the data and found it led to the same 76 items.

### Focus groups and Living/Lived Experience Input

To enhance face validity and ensure that MotDem reflected the living and lived experience of motivational change in later life, we used an iterative focus group approach as part of the item selection and refinement process. Three focus group sessions were conducted to determine which items best captured meaningful, real-world changes in motivation and pleasure, drawing on the 6 provisional domains identified during the panel review (Anticipatory, Effort/Persistence, Consummatory, Social, Satisfaction, Sensory).

**Focus Group 1** (N = 6) comprised research scientists, clinical neuropsychologists, neuropsychiatrists, and clinicians specialising in aging and dementia. Participants were presented with the candidate domains and corresponding items and were asked to evaluate, for each domain, which items best captured aspects of motivation and pleasure that may change early in healthy and pathological aging. Emphasis was placed on identifying items that captured subtle but clinically salient changes in motivation rather than mood-related symptoms. Across domains, items judged to most closely align with the domain construct and lived experience were prioritised, while items perceived as ambiguous, redundant, or overly abstract were discarded.

Participants then engaged in detailed review of item wording to ensure age-appropriateness, cultural accessibility, and educational fairness, with revisions made iteratively until consensus was reached. This process took approximately 2 hours and resulted in streamlining of the initial item pool from 76 to 40 items.

**Focus Group 2** (N = 7) comprised clinicians, research scientists, and trainees with specific expertise in dementia populations in which motivational and reward-processing changes are often prominent (e.g., Parkinson’s disease, frontotemporal dementia). Building on the outputs of Focus Group 1, participants reviewed the 40 shortlisted items with the aim of enhancing conceptual clarity and clinical relevance. As before, items were considered sequentially with closely related items combined to reduce redundancy. Attention was paid to refining item wording to maximise interpretability, including the removal of double negatives and abstract phrasing. Where appropriate, items were revised to include concrete and familiar examples of reward, to mirror how changes in motivation are typically described and recognised in daily life. As before, all items were evaluated for age– and cultural-appropriateness, and education-fairness, to ensure accessibility across diverse aging populations. This resulted in the item pool being further reduced from 40 to 28 items, retaining face validity while improving usability.

**Focus Group 3** (N = 8) comprised individuals with living and lived experience of dementia, including current and former care partners, and clinicians not directly involved in the study. The primary objective of this group was to establish face validity and ensure that the item pool captured motivational and hedonic changes that are recognisable in dementia populations. Participants were asked to consider whether the selected items reflected changes that would be noticeable in everyday life, whether the examples provided were age– and culturally-appropriate and without bias (e.g., socioeconomic), and whether any of the items felt ambiguous, inappropriate or missing.

This group reviewed the refined 28-item pool and endorsed the broad structure and content of the proposed questionnaire, with minor rewording to improve clarity and tone. Importantly, the panel identified additional aspects of motivation and pleasure that warrant consideration in the context of dementia but were not as well represented in the reduced item set. As a result, the panel requested the re-inclusion of 11 items originally prioritised by Focus Group 1. This process resulted in a final set of 39 unique items, to be taken forward for validation. The panel further reviewed response format and recall period, recommending a Likert-type scale indexing experiences over the past month to balance sensitivity with feasibility and acceptability for older adults and clinical populations.

### Validation of MotDem

The validation study (Stage 2) was conducted in March 2024 online via Qualtrics software on the MTurk platform and took approximately 40 minutes to complete. Following informed consent, participants completed the online Mini-ACE,^20^ which enabled us to screen for potential bots or automated responses. The prototype MotDem comprising 39 questions was completed, followed by validated questionnaires to probe related constructs of apathy, anhedonia, and mood (see below). For MotDem, participants were asked to consider each statement in relation to the past month and to indicate the frequency with which each item had occurred on a 4-point Likert scale (“Hardly Ever”, “Sometimes”, “Often”, “Almost Always”). Each item was scored from 1 to 4, with lower scores indicating greater impairment. Items 3, 24, 34 and 38 were reverse scored.

### Convergent validity of MotDem

Three standard scales were employed to establish the convergent validity of MotDem relative to the constructs of motivation and pleasure:

1) The Snaith-Hamilton Pleasure Scale (SHAPS)^21^, was used to index an individual’s self-reported capacity to experience pleasure. The SHAPS consists of 14 items with each rated on a 4-point Likert scale (“Strongly Disagree” to “Strongly Agree”), where lower scores indicate poorer hedonic tone (maximum score: 56).
2) Motivation was indexed using the Dimensional Apathy Scale (DApS)^22^, which contains three subscales measuring Executive, Emotional, and Behavioural/Cognitive initiation aspects of apathy. Items are rated on a 4-point Likert Scale (“Hardly Ever” to “Almost Always”) with higher scores denoting greater apathy (maximum score: 24 per subscale).
3) The depression subscale of the Depression Anxiety and Stress Scale (DASS-21)^23^ was included to assess the degree to which MotDem performance is associated with mood disturbances. This was motivated by the fact that, while broadly related, anhedonia and depression are dissociable in dementia populations.^24^ The DASS depression subscale (DASS-D) contains 7 items and is self-rated using a 4-point Likert scale (“Did not apply to me” to “Applied to me very much, or most of the time”) where higher scores denote greater depressive symptomatology (maximum score: 21).

## Statistical analyses

Bartlett’s test of Sphericity and the Kaiser–Meyer–Olkin test were used to evaluate the factorability of the data. Horn’s parallel analysis of principal factors was then used to determine the optimal number of factors for extraction.^25^ Then, an iterated principal axis exploratory factor analysis (EFA) was run using the Promax (oblique) rotation method with an a priori factor loading cut-off of ≥.40 and a cross-loading cut-off of ≥.32.^26^ An oblique rotation method was selected as factors were expected to be correlated.^26,27^ This step was repeated iteratively with the remaining items to determine the final factor structure. The internal consistency of the resulting questionnaire was evaluated by calculating average inter-item and item-scale correlations, and Cronbach’s alpha, for each factor and the total MotDem score. Convergent and discriminant validity of MotDem were assessed using Spearman’s Rho. *P* values were adjusted for multiple comparisons using the Bonferroni procedure with a critical alpha level of .05. Split-half reliability was determined by calculating Pearson’s correlation between odd and even numbered items. All procedures were performed in R (v4.5.3) using the *rstudioapi* (v0.18.0) and *psych* (v2.6.3) libraries. Descriptive statistics were performed in SPSS (v28.0) and hierarchical clustering for item-level visualisation was performed in Python (v3.12.5) using the SciPy library (v1.14.1).

## Normative modelling and cut-off scores

Normative modelling was performed on MotDem total scores to derive age-adjusted reference values for multidimensional motivation (Stage 3). Models were fitted in the discovery cohort using a Generalized Additive Model (GAM) to allow for potential non-linear variation in motivation across the adult lifespan. Models were implemented in the mgcv (v1.9.4), qgam (v2.0.0), and mgcViz (v0.2.1) libraries in R Studio (v2025.09.2+418). Model assumptions were evaluated using standard diagnostic procedures and influential observations were assessed and excluded if above established thresholds (i.e., >4/n).^28,29^ Age-adjusted normative ranges were derived from the fitted model, with cut-off values defined using a 90% inter-percentile range. Scores below the 10^th^ percentile were considered outside the normative range.

## Results

### Exploratory factor analysis (EFA)

To determine suitability of the data for EFA (Stage 2), the following screening was completed. Bartlett’s test of Sphericity (χ^2^[741] = 7402.67, *p* < .001) indicated that the correlation matrix was not an identity matrix, and the Kaiser-Meyer-Olkin statistic (.94) exceeded the recommended minimum (≥.70). Therefore, the 39-item questionnaire was considered appropriate for factor analysis and was submitted for EFA.

The initial EFA yielded a six-factor solution, as indicated by Horn’s parallel analysis, which accounted for 50% of the total variance. An iterative approach was adopted, resulting in four iterations, before arriving at a final solution, which consisted of 22 items with all loadings ≥.4 (see Supporting Information for revised questionnaire). Total scores on MotDem ranged from 22 to 88. Eigenvalues and the proportion of variance accounted for by each factor are presented in Table 1.

Each factor was assigned a label based on the theme across its respective items. Factor 1 was labelled “*Goal-Directed*”, reflecting motivated behaviour including initiation, effort expenditure and persistence (e.g., “I feel satisfied when I finish a task or learn a new skill”). Factor 2, named “*Social Rewards*”, captured the anticipation and enjoyment of activities that are social (e.g., “I enjoy receiving praise or compliments from other people”). Factor 3 was labelled “*Pleasure*”, and captured the hedonic experience, encompassing in-the-moment enjoyment of rewards (e.g., “I enjoy hearing pleasant sounds”). Factor 4, labelled “*Satiety*”, described satisfaction from task completion and efforts to prolong the pleasurable experience (e.g., “I like to prolong doing the things I enjoy for as long as possible”).

**Table 1.** Exploratory factor analysis solution for MotDem.

| **Factors** | **Label** | **Eigenvalue** | **Proportion variance** | **Cumulative variance** | **Item counts** |
| --- | --- | --- | --- | --- | --- |
| 1. | Goal-Directed | 7.61 | .16 | .16 | 7 |
| 2. | Social Rewards | 1.22 | .16 | .31 | 7 |
| 3. | Pleasure | 1.06 | .13 | .45 | 5 |
| 4. | Satiety | .88 | .04 | .49 | 3 |

Table 1 displays the four-factor solution from the principal axis exploratory factor analysis with Promax (oblique) rotation. We note that Satiety was below the common Eigenvalue cut-off of one but was included based on consensus.

### Internal consistency reliability

MotDem Total score displayed good internal consistency (Cronbach’s alpha, α = .90), as did three of the four MotDem subscales: *Goal-Directed*: α = .86; *Social Rewards*: α = .86; *Pleasure*: α = .83. Likewise, mean inter-item correlations were moderate for the *Goal-Directed, Social Rewards* and *Pleasure* subscales (*Goal-Directed*: Mean *r* = .48; *Social Rewards*: Mean *r* = .46; *Pleasure*: Mean *r* = .49), falling within recommended guidelines (i.e., .15 to .50)^30^. Finally, the average inter-item correlation for the overall measure was .30, with moderate item-scale correlations for the *Goal-Directed (r* = .63), *Social Rewards* (*r* = .63) and *Pleasure* (*r* = .62) subscales. The average item-scale correlation for the overall measure was moderate (*r* = .53) and split-half reliability of MotDem was strong (*r =*.93, *p*<.001). Overall, these metrics demonstrate good internal consistency reliability for MotDem Total and the 3 main factors of Goal-Directed, Social Rewards, and Pleasure.

The Satiety subscale did not demonstrate comparable psychometric properties, showing poor internal consistency (α =-.18) and weak inter-item (*r* =-.04) and inter-scale (*r* =-.06) correlations. Given this pattern, the Satiety subscale was treated as exploratory, an issue we return to below.

### Convergent validity: MotDem Total

MotDem Total demonstrated strong convergent validity with established measures of pleasure and motivation. Higher MotDem total scores were strongly associated with higher SHAPS scores (ρ = .72, *p* < .01) indicating greater hedonic experience (Figure 2.A). Similarly, MotDem Total showed a strong negative association with the DApS total (ρ = – .75, *p* < .01), such that higher motivation on MotDem was associated with lower levels of apathy.

**Figure 1.**
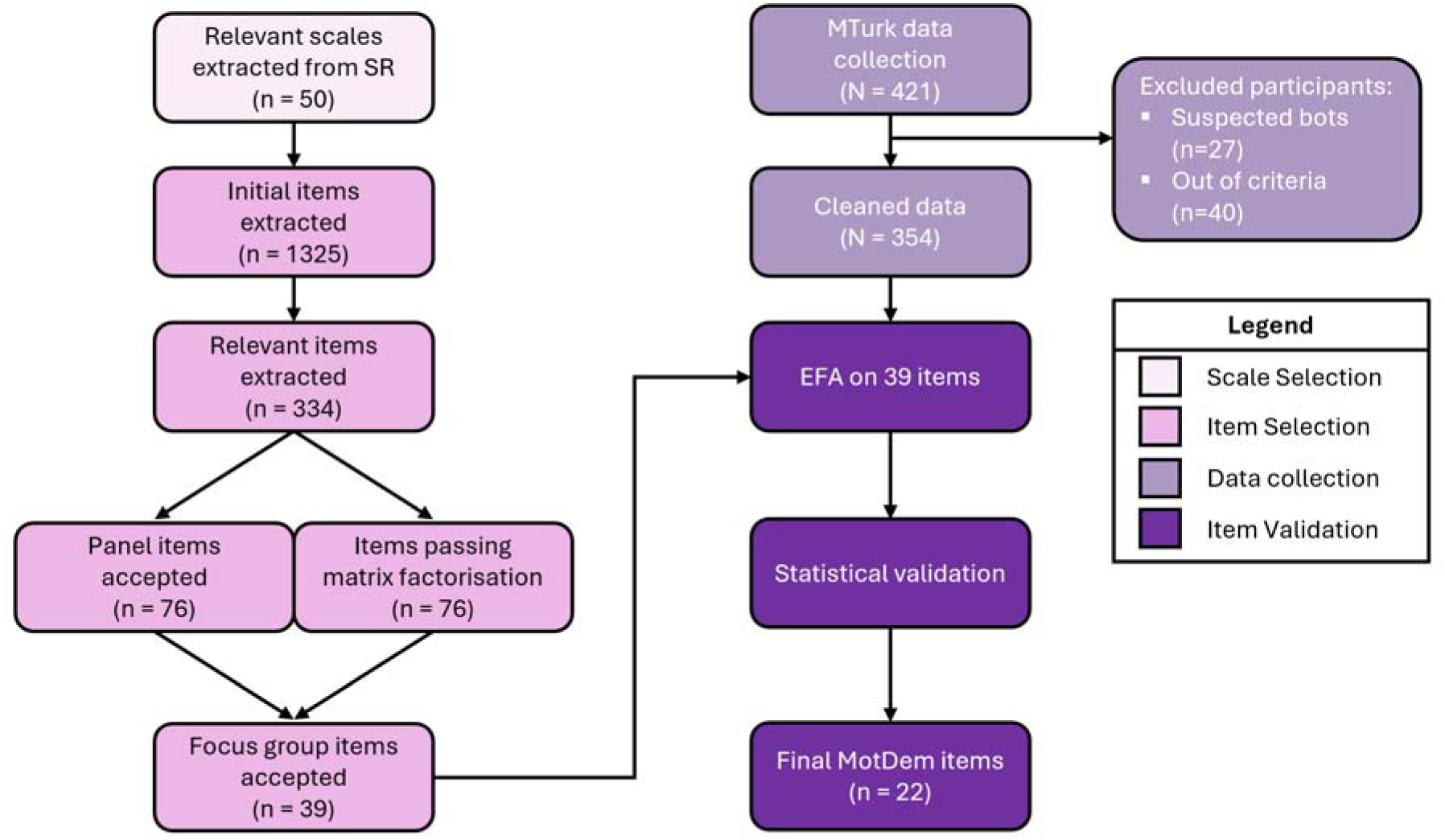
Overview of co-design, development, and validation process for MotDem. Abbreviations: SR = Systematic Review; EFA = Exploratory factor analysis; MTurk = Amazon Mechanical Turk.

**Figure 2.**
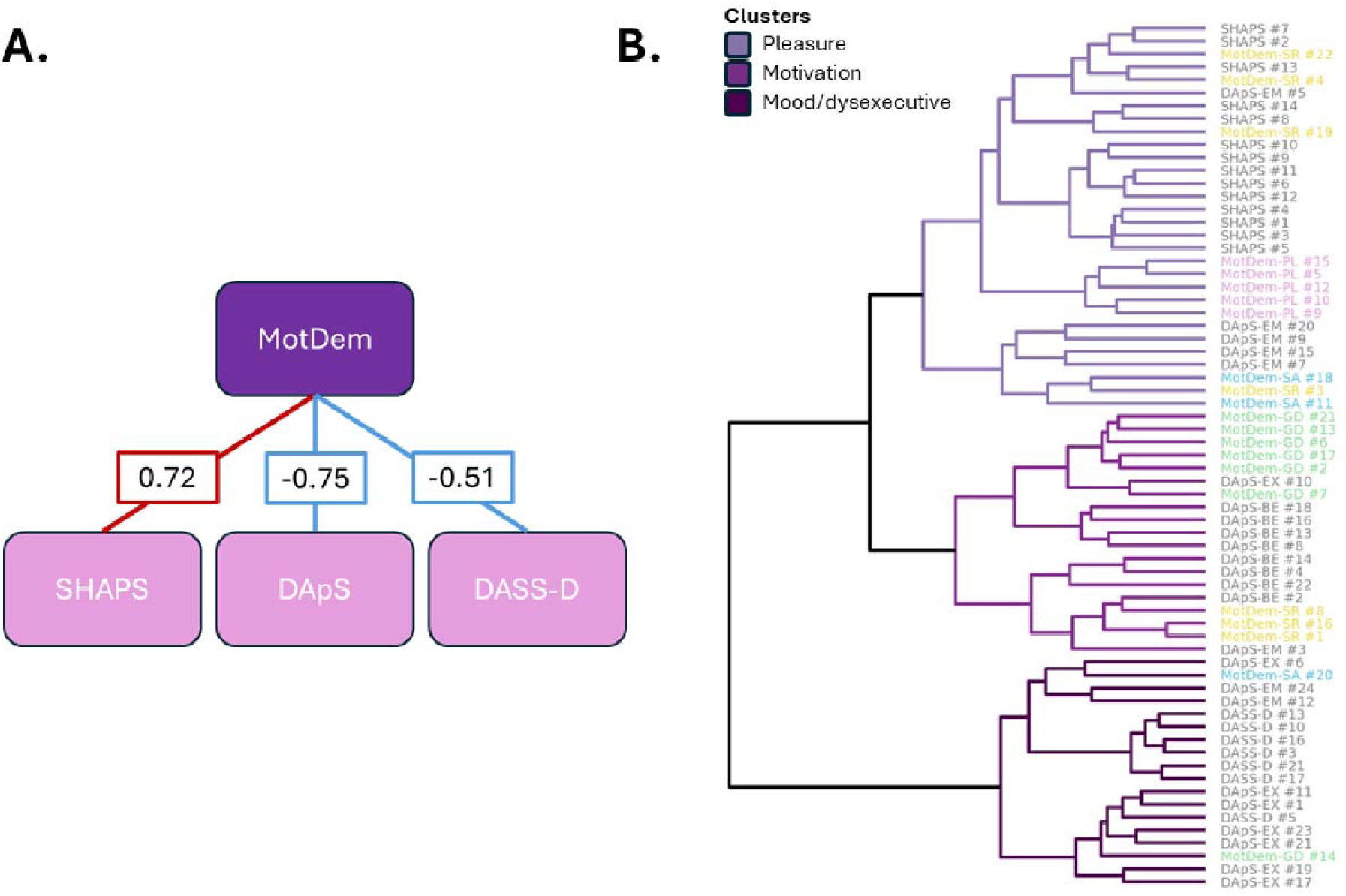
Multidimensional structure of MotDem and its relative dissociation from depressive symptomatology. Panel A depicts Spearman’s correlations between MotDem Total and related scales of pleasure (SHAPS), motivation (DApS), and mood (DASS-D). All *p* values remain significant after Bonferroni correction. Line colours denote negative (blue) or positive (red) associations. Panel B displays the hierarchical clustering performed on relevant items. Tree colours (i.e., purple shades) denote different construct clusters while MotDem items are coloured according to their subscales to improve visualisation. Abbreviations: DApS, Dimensional Apathy Scale; DASS-D, Depression Anxiety and Stress Scale Depression subscale; MotDem, Motivation in Aging and Dementia; SHAPS, Snaith-Hamilton Pleasure Scale.

By contrast, associations between MotDem Total and affective symptomatology were more modest. MotDem Total showed moderate negative correlations with the DASS-21 Total score (ρ = –0.47, *p* < .01) and the DASS-Depression subscale (ρ = –0.51, *p* < .01). This pattern indicates that while MotDem shares variance with affective disturbance, it more closely aligns with the target constructs of motivation and pleasure.

### Construct-level organisation of motivation indexed by MotDem

Hierarchical clustering was performed to examine item-level relationships across measures of motivation, pleasure and mood, including MotDem, SHAPS, DApS, and DASS-Depression subscale (Figure 2.B). Using Spearman’s correlation and Ward’s distance, three higher-order clusters emerged, broadly corresponding to pleasure, motivation, and mood/dysexecutive domains.

MotDem Pleasure and Satiety items clustered predominantly within the pleasure domain, alongside SHAPS items and the emotional apathy items from the DApS, indicating shared variance related to hedonic responsiveness. In contrast, MotDem Goal-directed items clustered with the DApS behavioural apathy items within the motivation domain, reflecting convergence on initiation, effort, and action-oriented aspects of motivation. MotDem Social Reward items showed a more distributed pattern, loading across both the pleasure and motivation clusters, consistent with the multidimensional and integrative nature of social reward processes.

The third cluster, characterised by mood and dysexecutive features, was dominated by the DASS-D items and executive apathy items from the DApS, with minimal representation from MotDem. This pattern indicates that MotDem items align more closely with motivational and hedonic constructs than with affective distress or dysexecutive symptoms.

### Convergent validity: MotDem subscales

Figure 3 shows associations between each MotDem subscale and corresponding measures from the DApS, SHAPS, and the DASS-Depression subscale. Within MotDem, subscales were inter-correlated to an acceptable degree, consistent with a multidimensional but related construct. The Pleasure, Social Reward, and Goal-Directed subscales showed moderate-to-strong intercorrelations (ρ *=* 0.55–0.63) indicating shared variance across core motivational dimensions.

**Figure 3.**
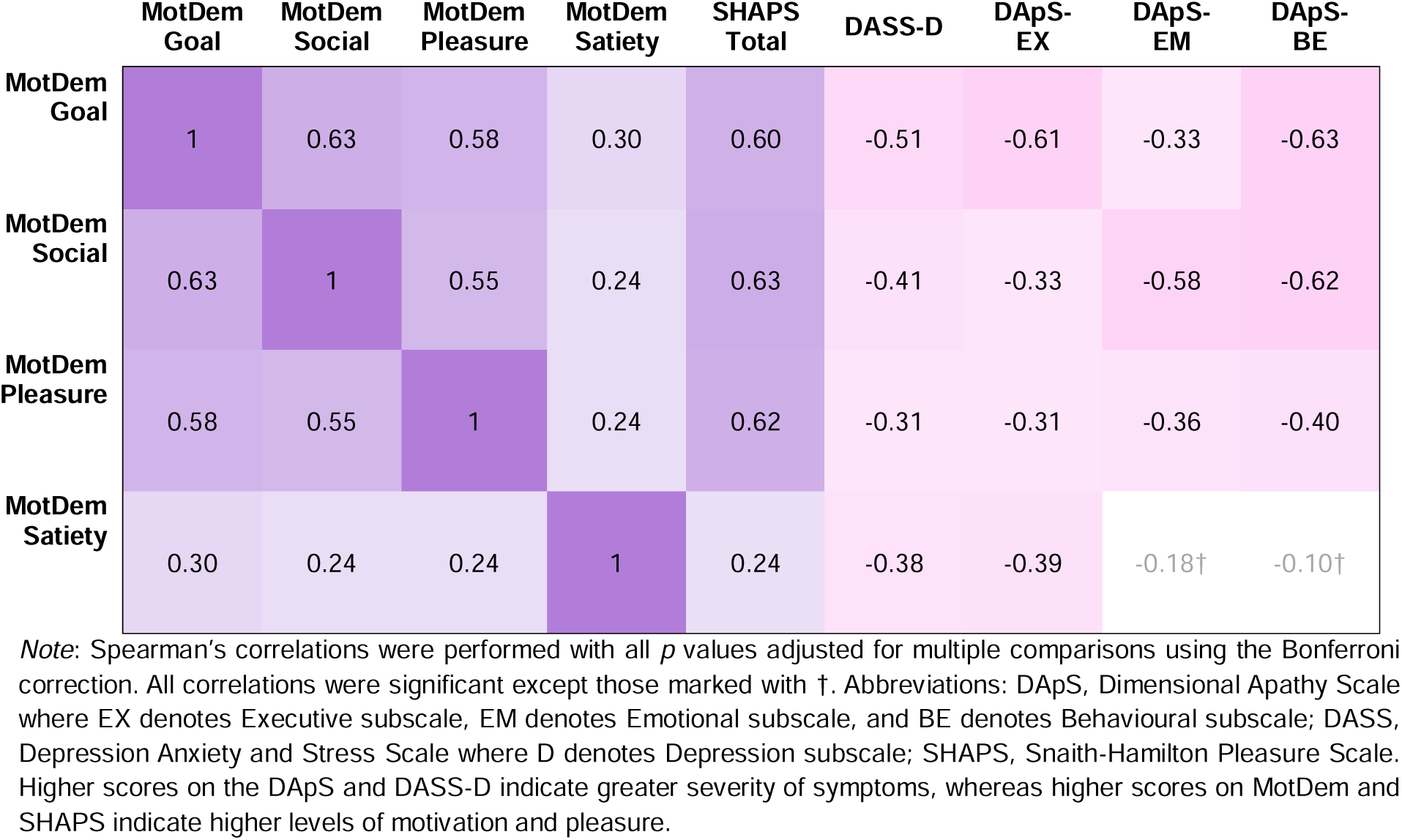
Correlation structure between MotDem subscales and external measures of apathy, anhedonia, and depression.

In contrast, the Satiety subscale showed weaker associations with the other MotDem subscales (ρ *=* 0.24–0.30). This pattern was anticipated given the distinct phenomenological focus of satiety-related items and their inclusion based on lived experience input to capture experiences relevant for pathological aging. Together, these findings support the internal coherence of the primary motivational dimensions while underscoring the exploratory nature of the Satiety subscale.

Across measures, all cross-scale correlations remained significant after Bonferroni correction (all *p* values < .01), except for associations between MotDem Satiety and the Behaviour and Emotion subscales of the DApS. Among MotDem dimensions, the Goal-Directed subscale showed the strongest convergence with apathy and reward-related measures, with robust negative associations with DApS Behaviour (ρ = –0.63), DApS Executive (ρ = –0.61), and SHAPS total score (ρ = 0.60). Similarly, the Social Reward subscale showed strong associations with both DApS Behaviour (ρ = –0.62) and SHAPS total score (ρ = 0.63) and was also related to the DApS Emotional subscale (ρ = –0.58). The Pleasure subscale demonstrated strong convergence with the SHAPS total score (ρ = 0.62) and moderate associations with DApS subdomains (ρ = –0.31 to –0.40).

The Satiety subscale showed the weakest convergence with established measures, with its highest associations observed with the DApS Executive (ρ = –0.39) and DASS-Depression (ρ = –0.38). Full correlation matrices, including weaker associations not discussed here (e.g., between DASS-Depression and MotDem subscales) are presented in Figure 3.

### External validation of MotDem in an older Australian sample

We next examined the robustness and cross-national generalisability of MotDem (i.e., the final 22 item version, Total score: 22-88) in the external validation cohort. Overall, MotDem demonstrated strong internal consistency in this older adult sample, closely mirroring the pattern observed in the US discovery cohort spanning the entire adult lifespan. Internal consistency indices were high for MotDem Total as well as Goal-Directed, Social Reward and Pleasure subscales (all α >.7), whereas the Satiety subscale again showed comparatively weaker reliability. Split half reliability for the total scale also remained strong (*r =*.90, *p*<.001). This convergent pattern across independent samples supports the stability of core motivational dimensions that are particularly salient in later life.

MotDem Total scores in the older Australian sample similarly showed good convergent validity, displaying moderate associations with established measures of anhedonia (SHAPS), apathy (DApS), and depressive symptoms (DASS-D). At the subscale level, convergent validity patterns were largely consistent with those observed in the US discovery sample. One exception was the emotional subscale of the DApS, which was not found to correlate with MotDem scores in the older Australian cohort. See Supplementary Material for full reliability and validity statistics.

### Age effects on motivational dimensions

We further examined age-related variation in MotDem scores across the broader discovery cohort spanning early to late adulthood (18-80 years). Age was a significant predictor of MotDem Total scores, accounting for a small proportion of variance (*F*(1, 352) = 21.81, *p* < .001, *R*^2^ = .06). Increasing age was associated with modestly higher MotDem scores [□ = 0.17, *t*(352) = 4.67, *p* <.001], reflecting age-related differences in motivational profiles across the adult lifespan. Importantly, the magnitude of this association was small, indicating that the core motivational dimensions indexed by MotDem remain relatively stable across adulthood (see Supplementary Material for further details).

### Normative modelling and online calculator

To enhance the interpretability and clinical utility of MotDem, we established age-adjusted normative ranges across the adult lifespan (Stage 3) using a generalised additive model (GAM). Age significantly predicted MotDem total scores [*F* = 18.62, *p* <.001], with a modest non-linear association across the lifespan (adjusted *r*^2^ = .09, deviance explained = 9.04%; Figure 4A), consistent with patterns observed in the linear analyses.

**Figure 4.**
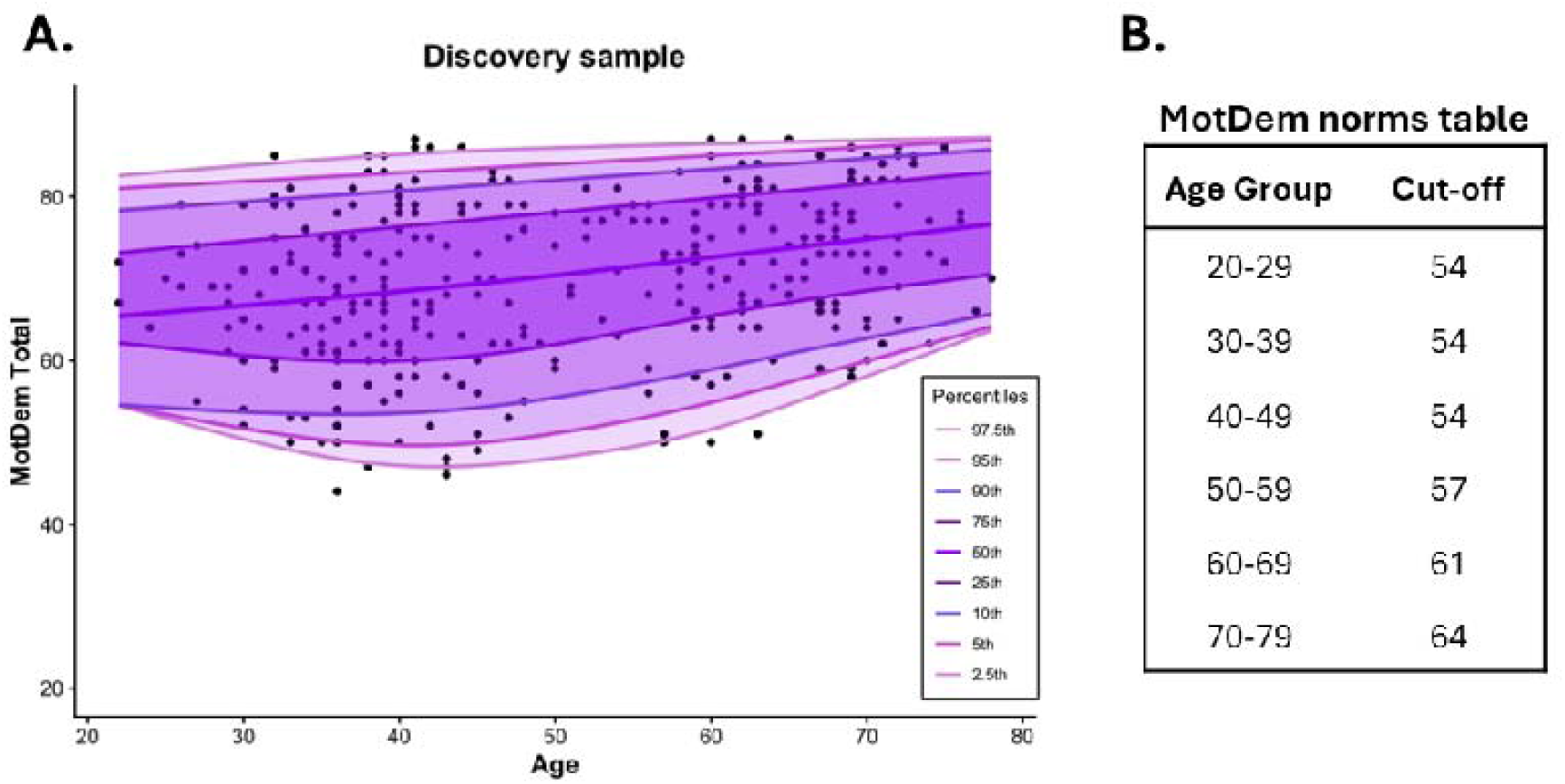
Normative model of MotDem Total and derived cut-off values. Panel A displays a normative model of MotDem (i.e., Motivation in Aging and Dementia) Total scores across age in the discovery sample (MotDem Total ranges from 22-88). Normative modelling was performed using a quantile-based GAM with cut-offs derived from the 10^th^ percentile and 90% inter-percentile range. Panel B depicts the derived cut-off values from the normative model across age bands.

Age-specific cut-off values were derived from the normative model using decade-based age bands to facilitate clinical application and account for sampling variation across the lifespan. Scores below the 10^th^ percentile-base were classified as falling outside the normative range (Figure 4.B, see Supplementary Table 3 for full quantile values). To support implementation, an online calculator is available to generate age-adjusted scores and cut-offs across MotDem subscales.

## Discussion

In this study, we introduce MotDem as a valid and reliable measure of multidimensional motivation and pleasure, co-designed for use across the adult lifespan and applicable to normative and clinical populations, including dementia. MotDem addresses a significant gap in existing assessment approaches by capturing distinct motivational domains (goal-directed behaviour, social reward, pleasure) that are not adequately represented in depression– or apathy-focused tools, yet are central to engagement, wellbeing, and aging trajectories. Crucially, the development of MotDem was embedded within a co-design framework involving people with lived experience of dementia, current and former carers, and clinicians, ensuring that the constructs identified through quantitative analysis reflect how motivational changes are experienced and described in everyday life. This alignment between data-driven structure and lived experience strengthens the face validity of MotDem and distinguishes it from existing tools that have tended to overlook socially salient or hedonic aspects of motivation.

Our findings support the view that motivation in aging is inherently multidimensional. Across two independent samples that differed in national context and age distribution, we identified and validated separable dimensions of Goal-Directed behaviour, Social Reward, and Pleasure. The emergence of these three distinct dimensions in both cohorts supports the presence of a common motivational architecture across adulthood. Rather than reflecting a cohort or context-specific effect, our findings suggest a stable and generalisable model of motivation in later life. This framework resonates with growing evidence that age-related motivational changes do not unfold uniformly but instead may vary across partially dissociable dimensions with distinct trajectories.^31,32^ Importantly, each of the three dimensions identified in MotDem displayed distinct patterns of association with external measures of apathy and anhedonia, underscoring their conceptual distinctiveness. Goal-Directed behaviour aligned most closely with behavioural and executive aspects of apathy, while Social Reward and Pleasure emerged as distinct factors separable from action-oriented motivation and depressive symptomatology.

The emergence of Social Reward as a standalone motivational dimension across the adult lifespan is noteworthy. Social motivation is commonly treated as a subdomain of apathy or as secondary to hedonic experience, however our findings indicate that social reward occupies a distinct position within the motivational architecture of aging. Across both samples, Social Reward was separable from Goal-Directed behaviour and showed only partial overlap with hedonic experience, suggesting that motivation to engage in and derive reward from social experiences reflects more than general action initiation or the capacity to experience pleasure. Sensitivity to social rewards likely encompasses the anticipatory valuation of social interactions, affective responsiveness, and socially motivated behaviour in ways that are not adequately captured by existing measures.^14^ This interpretation aligns with network-based perspectives in which certain psychological phenomena constitute distinct and replicable configurations of interrelated processes rather than dimensions of broader constructs.^33^ From this perspective, Social Reward may represent a motivational “natural kind” in aging, that is, a qualitatively distinct configuration rather than a simple subcomponent of apathy or anhedonia.

Pleasure also emerged as a distinct motivational dimension, resonating with mounting evidence that the hedonic experience is an important determinant of subjective wellbeing in later life.^9,10^ While pleasure is often subsumed within broader affective or depressive frameworks, our findings demonstrate that the hedonic experience shows only partial overlap with apathy and depressive symptomatology, reinforcing the importance of considering hedonic tone as a standalone dimension.^24^ This distinction appears to be salient in later life when the capacity to experience enjoyment may remain relatively preserved, or decline selectively, with such changes occurring independently of mood disturbances. While anticipatory valuation of reward is captured in the Social Reward construct, the Pleasure dimension appears to selectively capture consummatory hedonics, i.e., the in-the-moment experience of enjoyment. In older adulthood, consummatory pleasure may be crucial for emotional wellbeing and resilience, particularly in the context of health-promoting behaviours, mobility, and social engagement.^34^ In support of this view, pleasure has been shown to predict aging outcomes, including wellbeing and longevity, beyond the effects of health status, behavioural risk factors, and depressive symptoms.^9,10,35^ By explicitly indexing consummatory pleasure, MotDem provides a method of capturing motivational changes that are highly relevant for outcomes in later life, yet may go under-recognised in conventional assessments.^36^

Finally, it is important to note that a fourth motivational dimension relating to Satiety showed comparatively weaker psychometric properties across samples. This suggests that satiety-related experiences may not form a stable motivational dimension in healthy aging. It is important to contextualise the Satiety findings within the co-design process that led to their inclusion. Satiety-related changes, in relation to food, sex, and money, were highlighted by current and former care partners of people living with dementia as some of the most clinically salient and distressing symptoms for individuals and families. The complex relationship between reward processing and changes in satiety may limit the expression of such features in healthy or non-clinical samples, including the present validation cohort where individuals with current psychiatric diagnoses or significant psychotropic medication use were purposefully excluded. Importantly, alterations in eating behaviour^37,38^ and reward sensitivity^39,40^ have been documented as early features of several neurodegenerative disorders, suggesting that satiety-related processes may hold more clinical utility in pathological rather than healthy aging cohorts. Given that the next phase of this work will be to validate MotDem in clinical populations, including dementia, we elected to retain the Satiety dimension as exploratory. This will enable us to test whether satiety-related motivational changes emerge as meaningful features in neurodegenerative disorders, consistent with lived experience reports.

Collectively, these findings position motivation in aging as a multidimensional construct that is both empirically robust and grounded in lived experience. Social and hedonic aspects of motivation are garnering increasing attention in terms of their importance for wellbeing and sustained engagement in later life.^9,41^ Across independent samples spanning different age ranges and national contexts we demonstrate that Goal-Directed behaviour, Social Reward, and Pleasure represent conserved and conceptually distinct motivational dimensions, pointing to a generalisable motivational architecture across adulthood. Viewed in this light, age-related vulnerabilities are unlikely to arise from a global decline in motivation, but from selective shifts within specific motivational dimensions that differentially influence engagement, wellbeing, and long-term trajectories. Accordingly, global assessments of motivation in dementia may obscure targetable dimensions (e.g., consummatory pleasure) limiting opportunities to tailor interventions and support engagement. This has direct implications for rehabilitation and intervention design, where distinguishing between motivational dimensions may enable more targeted approaches, for example enhancing social engagement, supporting reward responsiveness, or facilitating goal initiation, rather than relying on global or non-specific strategies. Importantly, both social and hedonic forms of motivation have implications that extend beyond individual wellbeing. Promoting social motivation and engagement is not only essential for maintaining cognitive, emotional, and physical health in older adults but may also play a critical role in fostering more integrated and supportive communities across the lifespan.^16^ Considering how such shifts in motivation vary across different cultures and geographical regions will be an important next step for this work.

In conclusion, we present MotDem as a multidimensional assessment of motivation across the adult lifespan, developed through an iterative co-design process to ensure empirical robustness and lived-experience relevance. MotDem advances the field towards a consensus in how age-related motivational changes should be operationalised, and clarifies which motivational dimensions, namely Goal-Directed, Social Reward, and Pleasure, are essential for research and clinical assessment. Progress in understanding heterogeneous aging trajectories depends crucially on the ability to measure distinct motivational drivers rather than unitary or mood-centred assessments. In this context, MotDem provides a theoretically grounded and multidimensional addition to the aging assessment toolkit, offering a foundation for future longitudinal and mechanistic studies of resilience, vulnerability, and risk in later life.

## Data Availability

The final MotDem scale is provided in Supplementary Material. To ensure standardised administration and interpretation, the full testing pack, scoring materials and administration guide are available from the corresponding author (M. Irish).

## Acknowledgements

The authors thank the Sydney Dementia Network’s Lived Experience Expert Advisory Panel (SDN-LEEAP) for ongoing consultation in the development of MotDem. The authors also acknowledge the assistance of Peter Baldwin, Gina Llanes, Philip Mosley, and Lesley-Gaye Wong. This project is supported by a Medical Research Future Fund (MRFF) Dementia Aging and Aged Care grant to MI from the Australian Department of Health and Aged Care (GNT2024329). MI is supported by a National Health and Medical Research Council (NHMRC) of Australia Investigator Grant (GNT2025228). CX is supported by an Australian Research Council Future Fellowship (FT230100549). AG is supported through MRFF and NHMRC, the Dementia Australia Research Foundation, and philanthropic organisations. DF is supported by the Edwards Fund for Dementia Research. T. Chong is supported by grants from the Australian Research Council (FT220100294; DP250102224). S. Wong is supported by an NHMRC Investigator Grant (GNT1196904). OP is supported in part by an NHMRC Leadership Fellowship (GNT2008020). AW receives support from the NHMRC, MRFF, and the Wellcome Trust.

## Appendix

MotDem Consortium details:

Rebekah M. Ahmed: 0000-0001-6996-8317;^1,2^ Isabelle Burke: 0000-0003-0092-9760,^3^ Jade Cartwright: 0000-0002-6381-6184,^4^ Tao Chen: 0000-0002-4982-3951,^1,5^ Trevor Chong: 0000-0001-7764-3811,^6-8^ Sally Day: 0000-0002-1194-4360,^9^ David Foxe: 0000-0003-0299-3344,^1,5^ Fiona Kumfor: 0000-0002-3208-075X,^1,5^ Samantha Loi: 0000-0002-4953-4500,^11,12^ Olivier Piguet: 0000-0002-6696-1440,^1,5^ Siobhán Shaw: 0000-0002-4655-0898,^12,13^ Dennis Velakoulis: 0000-0002-8842-8479,^11^ Alexis E. Whitton: 0000-0002-7944-2172,^12,13^ Stephanie Wong: 0000-0002-0990-5039.^1,14^

^1^Brain & Mind Centre, The University of Sydney, Camperdown, NSW, Australia.

^2^Department of Neurology, Royal Prince Alfred Hospital, Sydney, NSW, Australia.

^2^School of Psychology, Deakin University, Burwood, VIC, Australia.

^4^School of Health Sciences, UTAS Health, University of Tasmania, Launceston, TAS, Australia.

^5^School of Psychology, The University of Sydney, Camperdown, NSW, Australia.

^6^Turner Institute for Brain and Mental Health, School of Psychological Sciences, Monash University, Clayton, Australia.

^7^Department of Neurology, Alfred Health, Melbourne, Australia.

^8^Department of Clinical Neurosciences, St Vincent’s Hospital, Melbourne, Australia.

^9^Sydney School of Heath Sciences, The University of Sydney, Camperdown, NSW, Australia.

^10^Department of Psychiatry, The University of Melbourne, Parkville, Victoria, Australia.

^11^Neuropsychiatry Centre, The Royal Melbourne Hospital, Parkville, Victoria, Australia.

^12^Black Dog Institute, University of New South Wales, Sydney, NSW, Australia.

^13^Faculty of Medicine and Health, University of New South Wales, Sydney, NSW, Australia.

^14^College of Human Sciences and Culture, Flinders University, Adelaide, SA, Australia

