## Supplementary for "Multidimensional motivation in aging: a validated framework spanning goal-directed behaviour, social reward and pleasure"

**Files included:**

**Supplementary Table 1.** Overview of questionnaires included in the item review for MotDem

**Supplementary Table 2.** Final MotDem item set following expert consensus and lived-experience input.

**Supplementary Figure 1.** Cross-national validation of MotDem total and subscales scores.

**Supplementary Figure 2.** Age-related variation in MotDem total scores across discovery and validation samples.

**Supplementary Figure 3.** Convergent validity of MotDem subscales in the older Australian validation cohort.

**Supplementary Table 3.** Percentile scores derived from the MotDem Total normative model.

**Supplementary Table 1.** Overview of questionnaires included in the item review for MotDem

| Scale | Reference |
| --- | --- |
| Alzheimer's Disease and Related Dementias Mood Scale | Tappen & Williams (2008) |
| Amotivational Syndrome Scale in Chronic THC Consumers | Castano et al. (2020) |
| Anhedonia Scale for Adolescents | Watson et al. (2021) |
| Anticipatory and Consummatory Eating Pleasure Scale | Bailly et al. (2020) |
| Anticipatory and Consummatory Interpersonal Pleasure Scale | Gooding & Pflum (2014) |
| Anxiety and Depression Questionnaire - Anhedonic Depression | Fajkowska et al. (2018) |
| Apathy Evaluation Scale | Marin et al. (1991) |
| Apathy Inventory | Robert et al. (2002) |
| Apathy Motivation Index | Ang et al. (2017) |
| Apathy Scale | Starkstein et al. (1992) |
| Asocial Beliefs Scale | Grant & Beck (2010) |
| Behavioural Inhibition System/Behavioural Activation System Scales | Carver & White (1994) |
| Chemosensory Pleasure Scale (Adult) | Zhao et al. (2019) |
| Demotivating Beliefs Inventory | Pillny et al. (2018) |
| Dimensional Anhedonia Scale | Rizvi et al. (2015) |
| Dimensional Apathy Scale | Radakovic & Abrahams (2014) |
| Domains of Pleasure Scale | Masselink et al. (2019) |
| Fawcett Clark Pleasure Scale | Fawcett et al. (1983) |
| Frontal Behavioral Inventory | Kertesz et al. (1997) |
| Frustrative Non-reward Responsiveness Scale | Wright et al. (2009) |
| Grit Scale | Duckworth et al. (2007) |
| Hedonic Deficit and Interference Scale | Frewen et al. (2012) |
| Homecare Measure of Engagement - Staff Questionnaire | Baker et al. (2016) |
| Hopelessness Depression Symptom Questionnaire | Metalsky & Joiner (1997) |
| Initiative-Interest Scale | Esposito et al. (2014) |
| Job Apathy Scale | Schmidt et al. (2017) |
| Lille Apathy Rating Scale | Sockeel et al. (2006) |
| Momentary Reports of Emotions and Pleasure Scale | Chentsova-Dutton et al. (2015) |
| Mood and Anxiety Symptoms Questionnaire - Short Form | Watson et al. (1995) |
| Motivation and Pleasure Scale – Self Report | Llerena et al. (2013) |
| Motive For Sensory Pleasure Scale | Eisenberger et al. (2010) |
| Multi-Dimensional Scale of Grit | Singh & Chukkali (2021) |
| Person-Environment Apathy Rating Scale | Jao et al. (2016) |
| Positive Valence System Scale | Khazanov et al. (2020) |
| Questionnaire for Eudaimonic Well-Being | Waterman et al. (2010) |
| Revised Physical Anhedonia Scale | Chapman & Chapman (1978) |
| Revised Social Anhedonia Questionnaire | Eckblad et al. (1982) |
| Rewarding Events Inventory | Hughes et al. (2017) |
| Schizotypal Traits Questionnaire for Young Adolescents - Revised Version | DiDuca & Joseph (1999) |
| Schizotypy Experimental Questionnaire^a^ | Venables & Rector (2000) |
| Self-Efficacy Scale | Sherer et al. (1982) |
| Self-Assessment Anhedonia Scale | Oliveres et al. (2005) |
| Self-Evaluation of Negative Symptoms | Dollfus et al. (2016) |
| Sensitivity to Punishment and Sensitivity to Reward Questionnaire | Torrubia et al. (2001) |
| Short O-LIFE Questionnaire | Mason et al. (2005) |
| Snaith-Hamilton Pleasure Scale | Snaith et al. (1995) |
| Specific Loss of Interest and Pleasure Scale | Winer et al. (2014) |
| Temporal Experience of Pleasure Scale - State Version (Modified) | Gard et al. (2006) |
| Thinking and Perceptual Style Questionnaire | Linscott & Knight (2004) |
| Vigour Assessment Scale | Dlagnekova et al. (2021) |

^a^Test name generated by PSYCHTests.

**Supplementary Table 2.** Final MotDem item set following expert consensus and lived-experience input.

| **Item** | **Question** |
| --- | --- |
| Q1 | I look forward to important events in my life (e.g., birthdays, family events, celebrations) |
| Q2 | I feel satisfied when I finish a task or learn a new skill |
| Q3 | I enjoy looking at pictures of people or things that are important to me (e.g., family, friends, pets) |
| Q4 | I get pleasure from helping others (e.g., doing favours, giving gifts, making food) |
| Q5 | I enjoy hearing pleasant sounds (e.g., birds chirping, laughter, ocean waves) |
| Q6 | I feel energised when I’m doing something I like |
| Q7 | Once I have started a task or activity, I like to see it through to the end |
| Q8 | I want to be around and connect with other people |
| Q9 | I find pleasure in small things (e.g., a bright sunny day, meeting a friend) |
| Q10 | I enjoy looking at beautiful things (e.g., a sunset, a starry night, scenery, art) |
| Q11 | I can't seem to get enough of some things (e.g., food, sex, money, music) |
| Q12 | I enjoy physical sensations (e.g., a warm bath, a refreshing shower, a deep stretch, a massage) |
| Q13 | I keep trying to achieve my goals even when there are setbacks |
| Q14 | I find it easy to get started on activities or tasks |
| Q15 | I enjoy smelling pleasant fragrances (e.g., freshy cut grass, flowers, fresh bread) |
| Q16 | I enjoy celebrating important occasions (e.g., birthdays, cultural/festive events) |
| Q17 | I enjoy satisfying my curiosity (e.g., learning new things, solving a problem) |
| Q18 | I like to prolong doing the things I enjoy for as long as possible |
| Q19 | I enjoy receiving praise or compliments from other people |
| Q20 | I get upset or angry if I am unable to do the things that I enjoy (e.g., hobbies, watching my TV show, going for a walk) |
| Q21 | I enjoy achieving my goals |
| Q22 | I enjoy spending time with people who care about me (e.g., family, close friends, carer, my pet) |

Colours represent the corresponding MotDem subscale of Goal-Directed Behaviour (green), Social Reward (yellow), Pleasure (pink), and Satiety (blue).

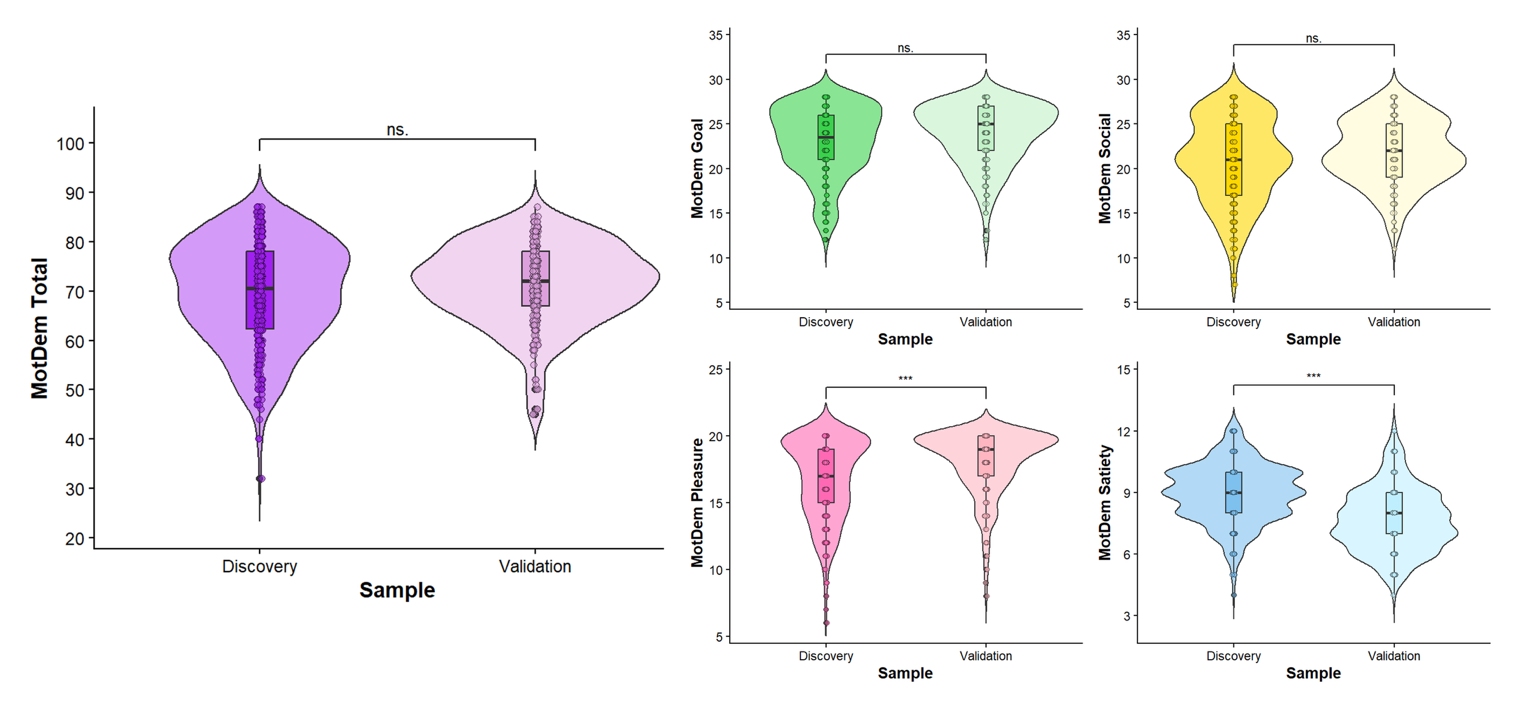
**Supplementary Figure 1.** Cross-national validation of MotDem total and subscales scores.

Distribution of MotDem total and subscale scores in the US discovery sample compared to the Australian validation cohort. Left panel displays MotDem total scores (purple), while right panels display Goal-Directed (green), Social Reward (yellow), Pleasure (pink), and Satiety (blue) subscale scores. Higher scores reflect greater levels of motivation. MotDem Total Max score = 88. Wilcoxon rank sum tests were run to compare group differences between discovery (MTurk) and validation (StepUp) samples on MotDem total and each of the four subscales, with Bonferroni correction applied (corrected *p* value = 0.01). No significant differences were observed for MotDem Total scores (*W* = 39166, *p* = .09), Goal-Directed (*W* = 37908, *p* = .021) or Social Reward (*W* = 38310, *p* = .034), however, a significant group effect was found for Pleasure (*W* = 32138, *p <* .001) and Satiety (*W* = 62953, = *p <* .001) subscales, reflecting higher levels of pleasure and lower levels of satiety in the older validation group.

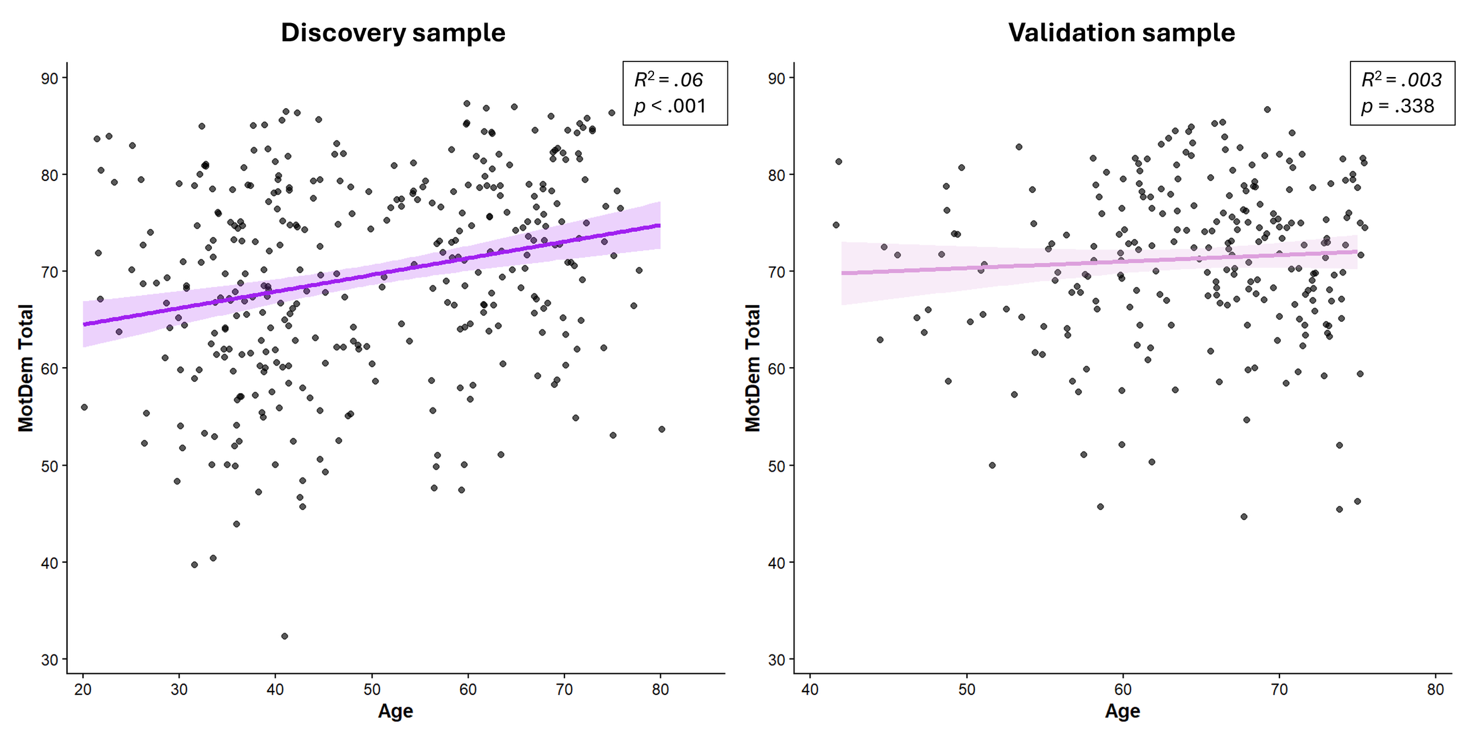
**Supplementary Figure 2.** Age-related variation in MotDem Total scores across discovery and validation samples.

Linear regressions with age as a predictor of MotDem Total in the US discovery sample (left) and Australian validation cohort (right). A modest positive association was observed in the discovery sample [ꞵ = 0.17, *t*(352) = 4.67, *p* <.001] with a small accounted variance (*R*^2^ = .06), whereas no significant relationship was observed in the older validation cohort (*p* = .338). This pattern indicates relative stability of MotDem scores in healthy older adults, alongside modest age-related variation across the broader adult lifespan.

**Supplementary Figure 3.** Convergent validity of MotDem subscales in the older Australian validation cohort.

|  | **MotDem Goal** | **MotDem Social** | **MotDem Pleasure** | **MotDem Satiety** | **SHAPS Total** | **DASS-D** | **DApS-EX** | **DApS-EM** | **DApS-BE** |
| --- | --- | --- | --- | --- | --- | --- | --- | --- | --- |
| **MotDem Goal** | 1 | 0.71 | 0.61 | 0.62 | 0.50 | -0.58 | -0.46 | -0.01† | -0.50 |
| **MotDem Social** | 0.71 | 1 | 0.49 | 0.60 | 0.47 | -0.43 | -0.30 | -0.11† | -0.36 |
| **MotDem Pleasure** | 0.61 | 0.49 | 1 | 0.58 | 0.53 | -0.47 | -0.36 | -0.11† | -0.46 |
| **MotDem Satiety** | 0.62 | 0.60 | 0.58 | 1 | 0.48 | -0.48 | -0.32 | -0.14† | -0.47 |

*Note*: Spearman’s correlations performed with all *p* values adjusted for multiple comparisons using Bonferroni correction. All correlations were significant except those marked with †. Abbreviations: DApS, Dimensional Apathy Scale where EX denotes Executive subscale, EM denotes Emotional subscale, and BE denotes Behavioural subscale; DASS, Depression Anxiety and Stress Scale where D denotes Depression subscale; SHAPS, Snaith-Hamilton Pleasure Scale. Higher scores on the DApS and DASS-D indicate greater severity of symptoms, whereas higher scores on MotDem and SHAPS indicate higher levels of motivation and pleasure.

The Australian older population showed strong internal consistency (i.e., *α*), mean inter-item correlation (i.e., *r_item_*), and item-scale correlation (i.e., *r_scale_*) across MotDem Total (*α* = .85, *r_item_* = .21 , *r_scale_* = .42), Goal-Directed (*α* = .83, *r_item_* = .43, *r_scale_* = .59), Social Rewards (*α* = .74, *r_item_* = .29, *r_scale_* = .46), and Pleasure (*α* = .77, *r_item_* = .41, *r_scale_* = .55) subscales. The Satiety subscale continued to display relatively weaker results (*α* = -.30, *r_item_* = -.07, *r_scale_* = -0.10).

In terms of convergent validity, MotDem Total scores in the Australian older cohort displayed moderate associations with established measures of anhedonia (SHAPS; *ρ* = 0.59, *p* < .001), apathy (DApS; *ρ* = -0.55, *p* < .001), and depressive symptoms (DASS-D; *ρ* = -0.58, *p* < .001). For the individual subscales, the only difference to emerge from the US was that the DApS emotional subscale was not significantly associated with any of the MotDem metrics in the older Australian sample (all *ρ's* = -.01 to -.14 and all *p*'s > 0.04 with corrected α = .0015).

**Supplementary Table 3.** Percentile scores derived from the MotDem Total normative model.

| Age | 2.5th Percentile | 5th Percentile | 10th Percentile | 25th Percentile | 50th Percentile | 75th Percentile | 90th Percentile | 95th Percentile | 97.5th Percentile |
| --- | --- | --- | --- | --- | --- | --- | --- | --- | --- |
| 20-29 | 52 | 53 | 54 | 61 | 66 | 74 | 79 | 81 | 83 |
| 30-39 | 49 | 51 | 54 | 60 | 67 | 75 | 80 | 82 | 84 |
| 40-49 | 47 | 50 | 54 | 61 | 69 | 77 | 81 | 83 | 86 |
| 50-59 | 49 | 53 | 57 | 63 | 71 | 79 | 83 | 84 | 86 |
| 60-69 | 54 | 57 | 61 | 67 | 74 | 81 | 84 | 85 | 87 |
| 70-79 | 61 | 62 | 64 | 69 | 76 | 82 | 85 | 86 | 87 |

MotDem Total scores range from 22 to 88. Percentile values we extracted from the normative model for each age represented, then averaged across each decade to ensure ease of interpretation and robustness. Values were rounded to the nearest full number to correspond with the format of MotDem scores. Ages at each end of the spectrum can sometimes lack representation and values should be interpreted with this in mind.
